# Corrected Visual Acuity as the Foundation of Effective Myopia Control: A 1-Year Real-World Cohort Study in 9-Year-Old Children

**DOI:** 10.64898/2026.03.17.26348596

**Authors:** You-Ruo Zhang, Qiu-Lin Mi, Huan Xiao, Ying-Ying Nie, Yi-Chun Chai, Ting Li, Junguo Duan

**Affiliations:** School of Ophthalmology, Chengdu University of Traditional Chinese Medicine, Chengdu, China; Sichuan Provincial Key Laboratory of Traditional Chinese Medicine Eye Health, Chengdu, China; Independent Researcher

**Keywords:** Myopia control, Corrected visual acuity, Axial length elongation, School-aged children, Single-vision spectacles, Real-world study, Propensity score matching

## Abstract

To determine whether achieving normal corrected visual acuity independently influences myopia progression in school-aged children wearing single-vision spectacles. In a one-year real-world cohort study, 9-year-old myopic children were classified into three groups: uncorrected, adequately corrected (normal visual acuity), and under-corrected (subnormal visual acuity). One-to-one propensity score matching was used to balance baseline characteristics, and annual axial length growth was compared. The adequately corrected group showed the slowest axial elongation (0.23 ± 0.14 mm/year), significantly less than both the under-corrected (0.35 ± 0.14 mm/year) and uncorrected groups (0.37 ± 0.16 mm/year) (all P < 0.001). Although the under-corrected group exhibited marginally slower progression than the uncorrected group, this minimal benefit was not sustained in semiannual analyses and lacked clinical relevance. Simply prescribing spectacles is insufficient for myopia control; achieving normal corrected visual acuity is essential to meaningfully slow axial elongation.

## Introduction

Myopia progression in children constitutes a critical global public health challenge, with early-onset cases substantially elevating the lifetime risk of high myopia and vision-threatening complications such as myopic maculopathy and retinal detachment[1–3]. Although spectacles represent the most prevalent first-line optical intervention[4,5], their efficacy in mitigating myopia progression depends not merely on refractive prescription accuracy but critically on achieving and maintaining optimal corrected distance visual acuity (CDVA)[6–8]. A large-scale epidemiological survey of myopia among children aged 6–18 years in Chengdu, China, reported a spectacle-wearing prevalence of 65.7%; however, 50.5% of spectacle wearers exhibited undercorrection despite lens use[9]. This persistent suboptimal CDVA sustains retinal defocus, thereby accelerating axial elongation[10–12]. Nevertheless, prior longitudinal studies have failed to isolate visual acuity status—specifically, persistently suboptimal best-corrected distance visual acuity (<0.0 logMAR) or persistently suboptimal uncorrected distance visual acuity—as an independent exposure variable[13–15]. Ethical constraints preclude randomized allocation of children to undercorrected conditions; consequently, high-quality observational studies employing robust causal inference methodologies represent the highest feasible evidence level for addressing this clinical question. This 1-year real-world cohort study enrolled 9-year-old children using only single-vision spectacles without adjunctive myopia control interventions (e.g., low-dose atropine, orthokeratology, or specialized myopia-control lenses). Using propensity score matching to minimize confounding bias, we aimed to determine whether children with persistently suboptimal CDVA (<0.0 logMAR) demonstrated significantly faster annualized axial elongation compared to those achieving target CDVA (≥0.0 logMAR) or uncorrected peers. The central hypothesis posits that the quality of visual rehabilitation—not merely spectacle provision—constitutes the cornerstone of effective myopia control.

## Methods

### Study Design and Ethical Compliance

This investigation was a school-based retrospective cohort study with longitudinal follow-up, utilizing vision screening data from a citywide childhood myopia prevention program in Chengdu, Sichuan Province, China (2019–2024). The study adhered to the tenets of the Declaration of Helsinki. Ethical approval was granted by the Ethics Committee of the Affiliated Eye Hospital of Chengdu University of Traditional Chinese Medicine (Approval No.: 2019YH-007). Written informed consent was obtained from parents or legal guardians of all participants through collaboration with the Chengdu Municipal Education Bureau.

### Study Participants and Eligibility Criteria

Eligible participants were children aged 9-year-old with myopia defined as spherical equivalent (SE) between –0.50 D and –6.00 D. Inclusion required: (1) complete baseline data for age, sex, uncorrected visual acuity (UCVA), corrected distance visual acuity (CDVA), SE, axial length (AL), anterior chamber depth (ACD), lens thickness (LT), and corneal curvature; (2) ≥3 consecutive follow-up visits over 1 year; (3) no prior myopia intervention or exclusive use of single-vision spectacles (no atropine, orthokeratology, or specialized myopia-control lenses); and (4) absence of data inconsistencies. Exclusion criteria included organic ocular disease, anisometropia >1.50 D, or cognitive impairment affecting visual testing reliability.

### Ophthalmic Examination Protocol

All examinations were performed on school premises by certified optometrists at 6-month intervals. UCVA was measured at 5 m using the standardized logarithmic visual acuity chart (GB 11533) under illumination >500 lux. Spectacle wearers underwent additional CDVA assessment, and spectacle type was recorded. Anterior segment evaluation used slit-lamp biomicroscopy. Non-cycloplegic autorefraction was conducted using Topcon KR-1 (Topcon Corporation, Tokyo, Japan) or Nidek ARK-1 (Nidek Co., Ltd., Gamagori, Japan) with triplicate measurements per eye; if any two readings differed by >0.50 D, additional triplets were taken until intra-measurement variation was <0.50 D. SE was calculated as sphere + ½ cylinder. Concurrently, axial length (AL), corneal curvature (K1, K2), ACD, and LT were measured using the Sovision SW-9000 optical biometer (Sovision Medical Technology Co., Ltd., Chongqing, China). AL readings differing by >0.02 mm were repeated. All devices underwent daily calibration; 10% of samples were independently re-measured by two operators for quality control.

### Outcome Definitions and Group Classification

Participants were classified into three mutually exclusive groups based on 1-year longitudinal data:(1) Untreated Screened Myopia Group (USMG): Myopic children (SE ≤ –0.50 D, UCVA >0.0 logMAR) without any optical correction during follow-up;(2) Protocol-Defined Optimal Correction Group (POCG): Consistent single-vision spectacle wearers with CDVA <0.0 logMAR at all visits;(3) Under-Corrected Group (UCG): Consistent single-vision spectacle wearers with CDVA ≥0.0 logMAR at all visits.

All groups received no adjunctive interventions. The primary outcome was annualized axial length change (ΔAL, mm/year) over 1 years. Secondary outcomes included first-time(ΔAL1) and second-time(ΔAL2) elongation rates.

### Statistical Analysis

Analyses used right-eye data only to avoid inter-eye correlation. Data management and statistics were performed in Python 3.9.6 (Statsmodels 0.14.5). Continuous variables (mean ± SD) were compared using one-way ANOVA; categorical variables (%) used chi-square tests. Propensity scores were estimated via logistic regression adjusting for age, sex, baseline SE, AL, K1, K2, ACD, and LT. Matching was stratified by sex (exact) and SE (<–3.00 D; –3.00 to –6.00 D), using 1:1 nearest-neighbor matching (caliper = 0.2 × SD of logit propensity score). Post-matching balance was assessed by absolute standardized mean differences (|SMD|<0.1 indicating balance). ΔAL comparisons in matched cohorts used paired t-tests. Results are reported as mean differences with 95% confidence intervals; P<0.05 (two-tailed) indicated significance.

### Participant Flow

The participant selection and analytical workflow are illustrated in a CONSORT-style flow diagram (Figure 1).

**Figure 1.**
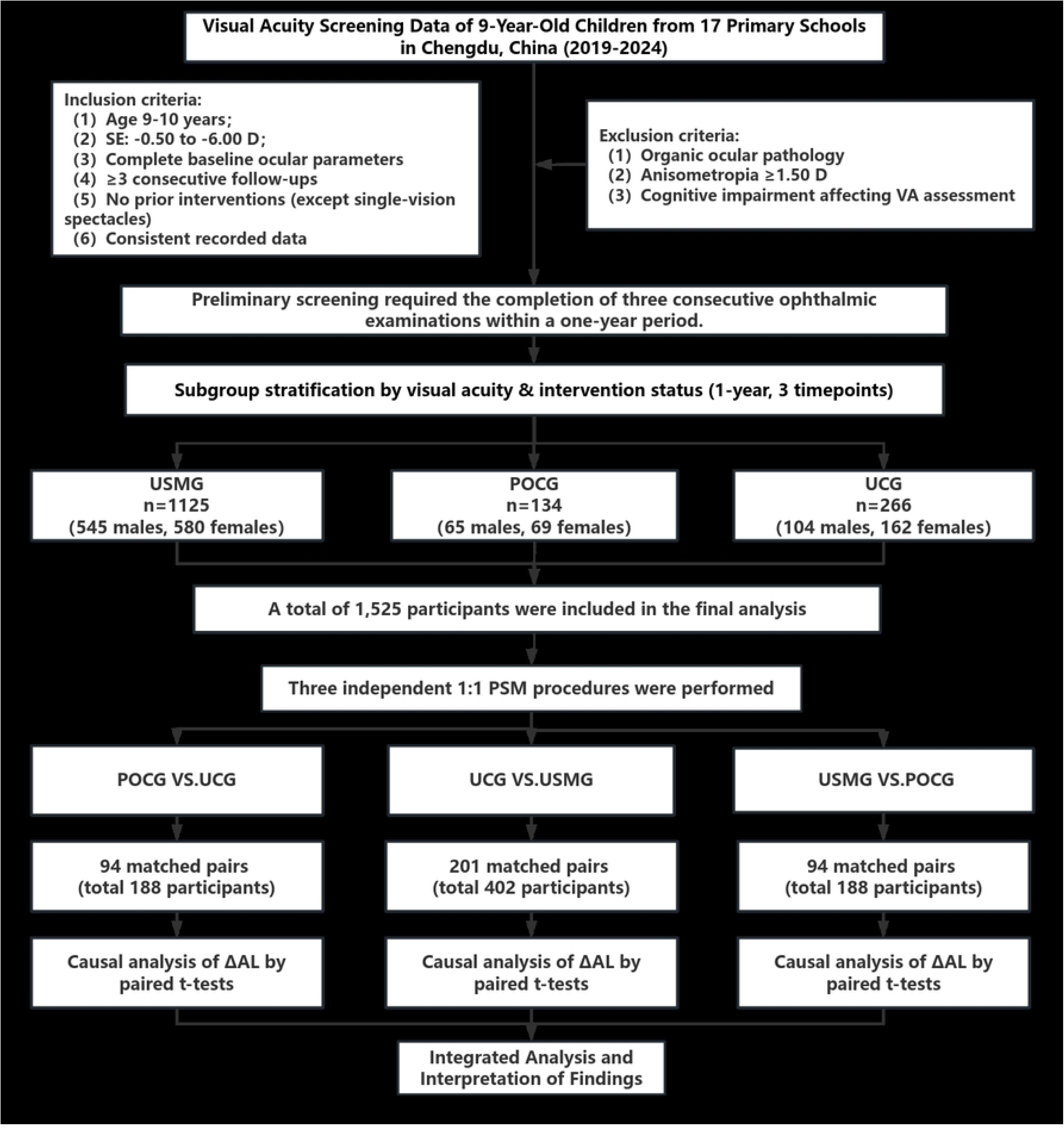
CONSORT flow diagram of participant selection, group allocation, and analytical pathways.

POCG,Protocol-Defined Optimal Correction Group; USMG,Untreated Screened Myopia Group; UCG,Under-Corrected Group.

## Results

### Study Population

As shown in Table 1 and Figure 1, this retrospective cohort study included 1,525 children aged 9 years at baseline, all with complete visual function and intervention status data across three consecutive annual examinations. Participants were classified into three groups: untreated screened myopia group (USMG, n=1,125), protocol-defined optimal correction group (POCG, n=134), and undercorrection group (UCG, n=266). Baseline demographic and ocular biometric characteristics are summarized in Table 1.

**Table 1.**
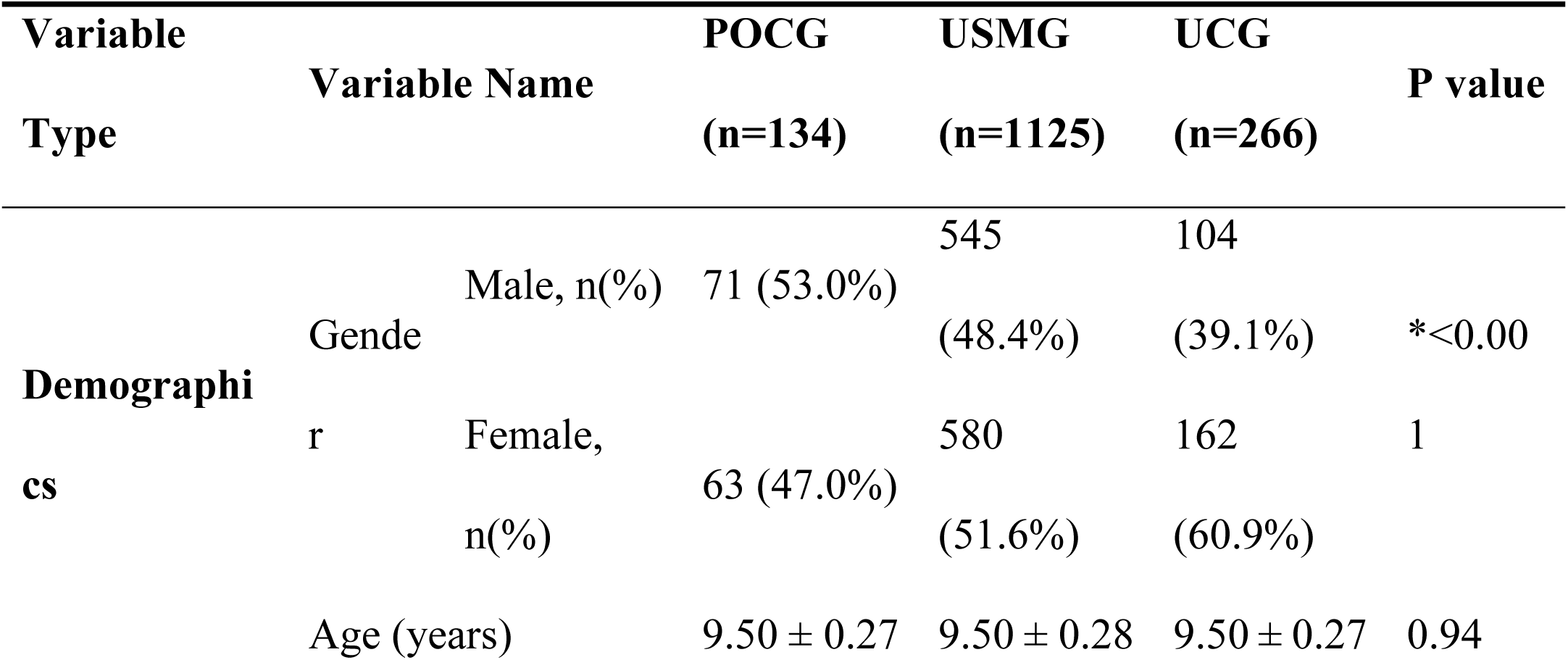
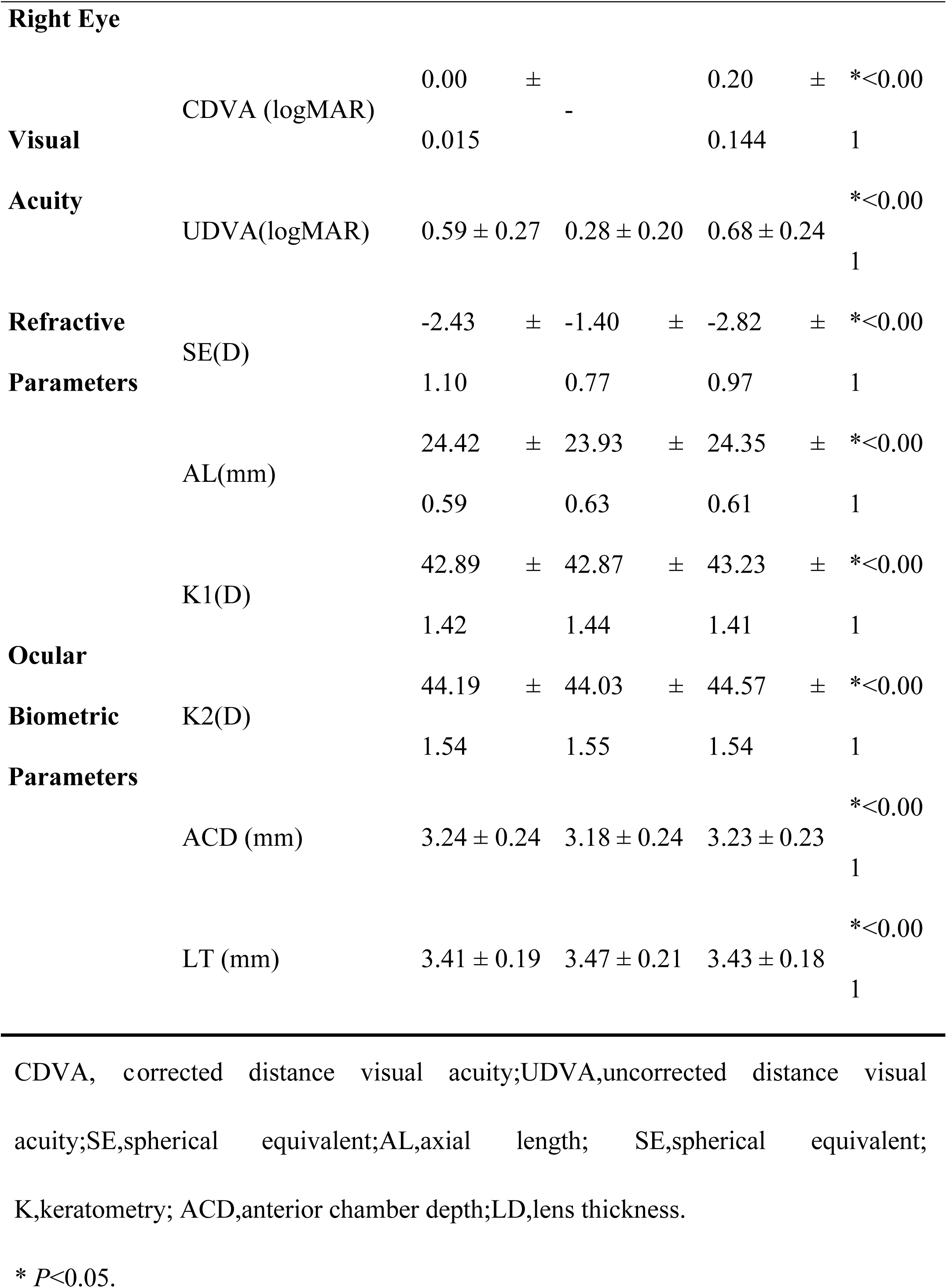
Demographic characteristics of participants across refractive correction status groups prior to propensity score matching.

### Propensity Score Matching Results

To address baseline imbalances, three independent 1:1 propensity score matching (PSM) procedures were performed, with exact matching on sex and stratification by baseline spherical equivalent (<–3.00 D; –3.00 to –6.00 D). Standardized mean differences (SMDs) before and after matching are reported in Tables 2–4 and visualized in Love plots (Figures 2–4).

**Figure 2.**
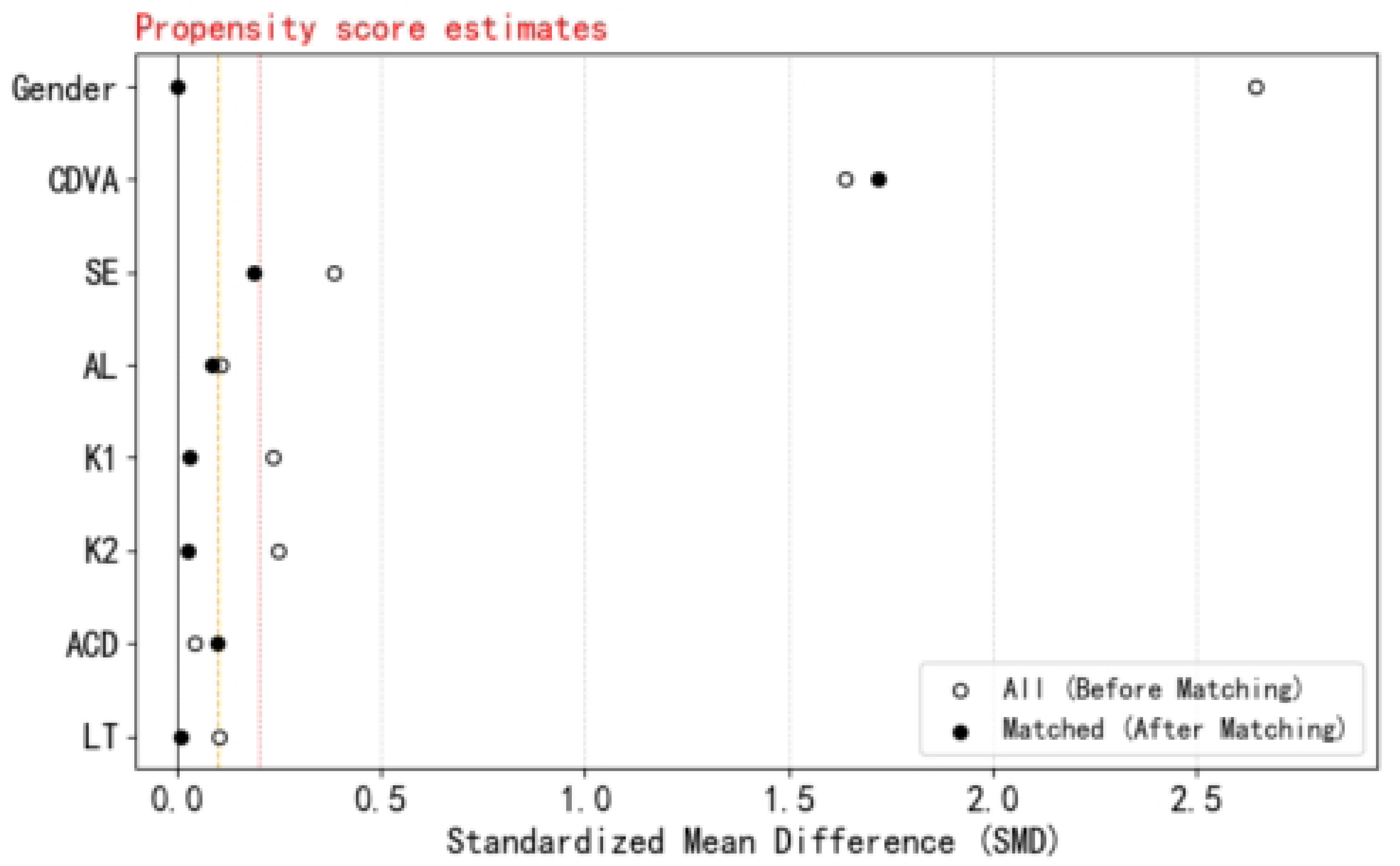
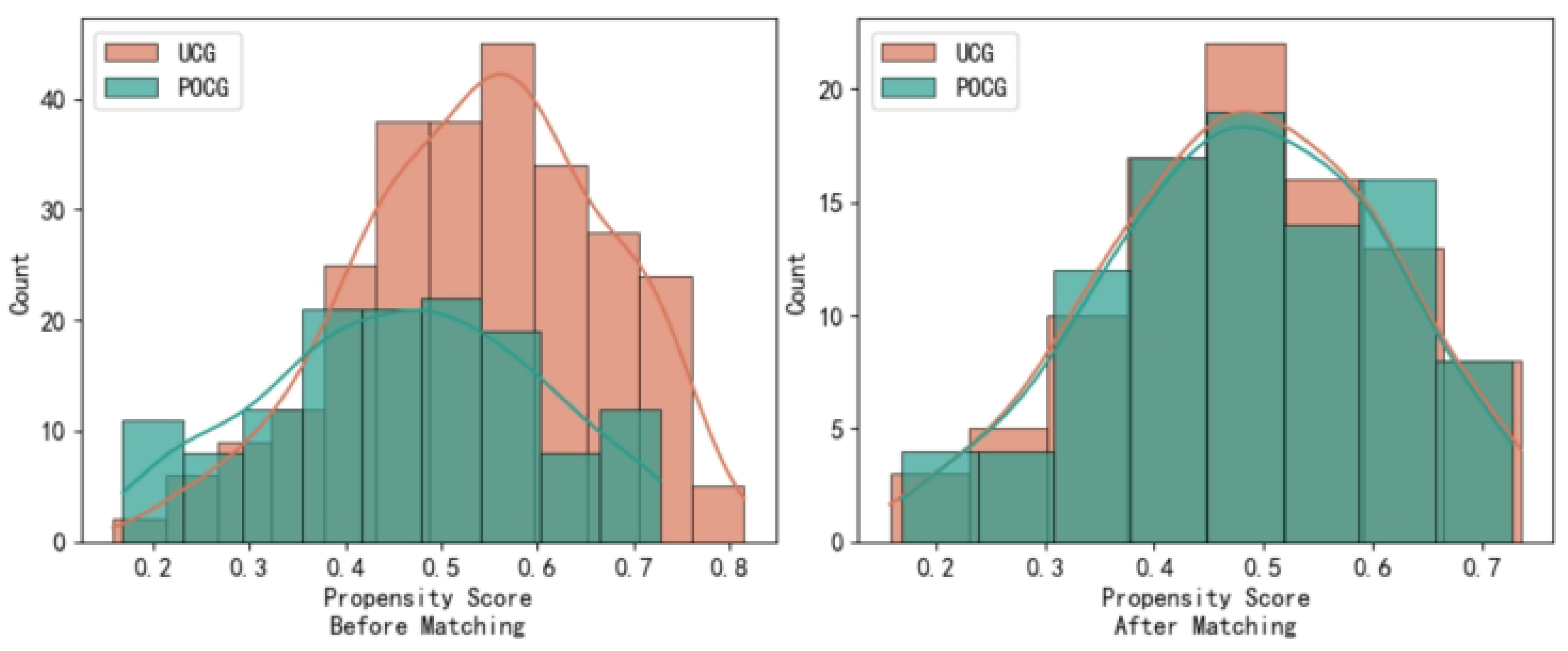
Love Plot and Distribution Fitting Curves for Baseline Characteristics Before and After Propensity Score Matching Between UCG and POCG.

**Figure 3.**
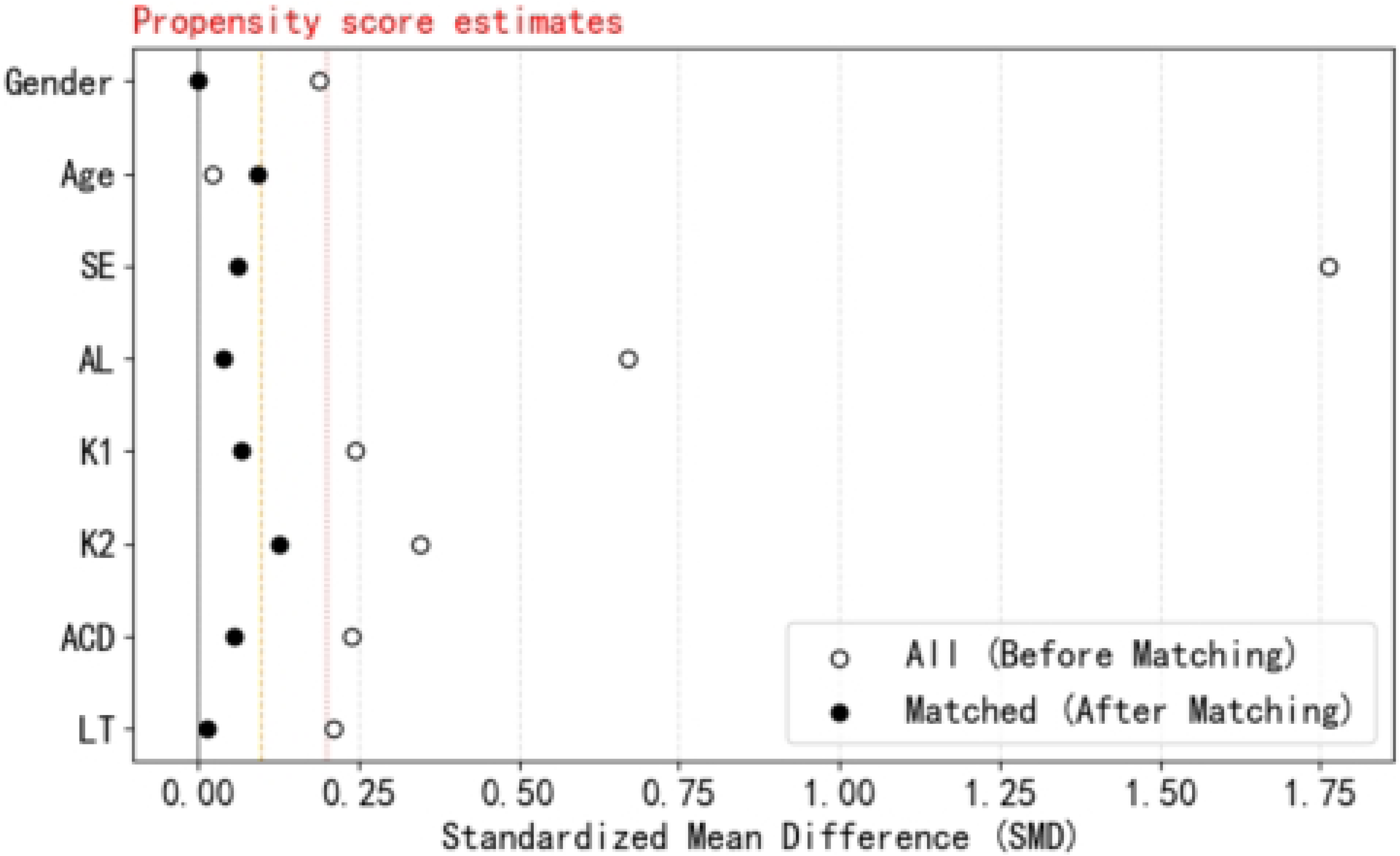
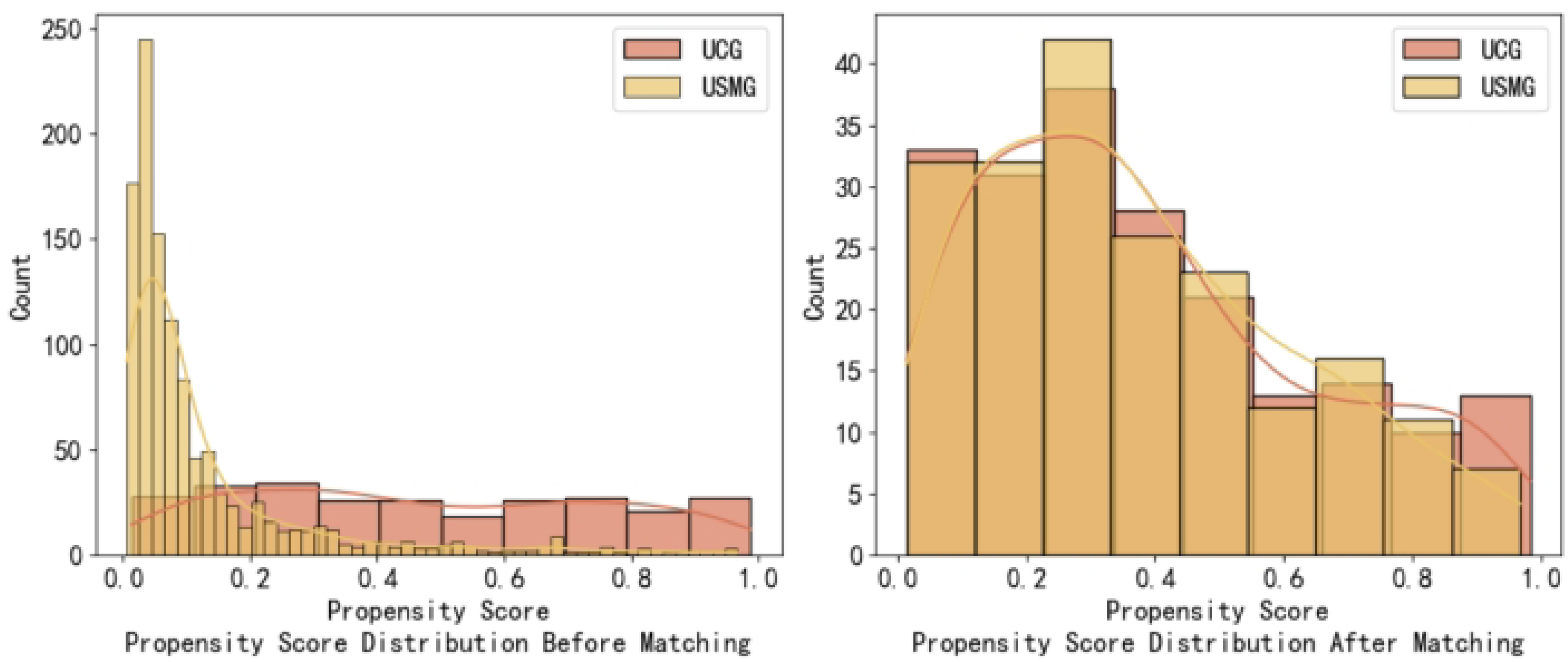
Love Plot and Distribution Fitting Curves for Baseline Characteristics Before and After Propensity Score Matching Between UCG and USMG.

**Figure 4.**
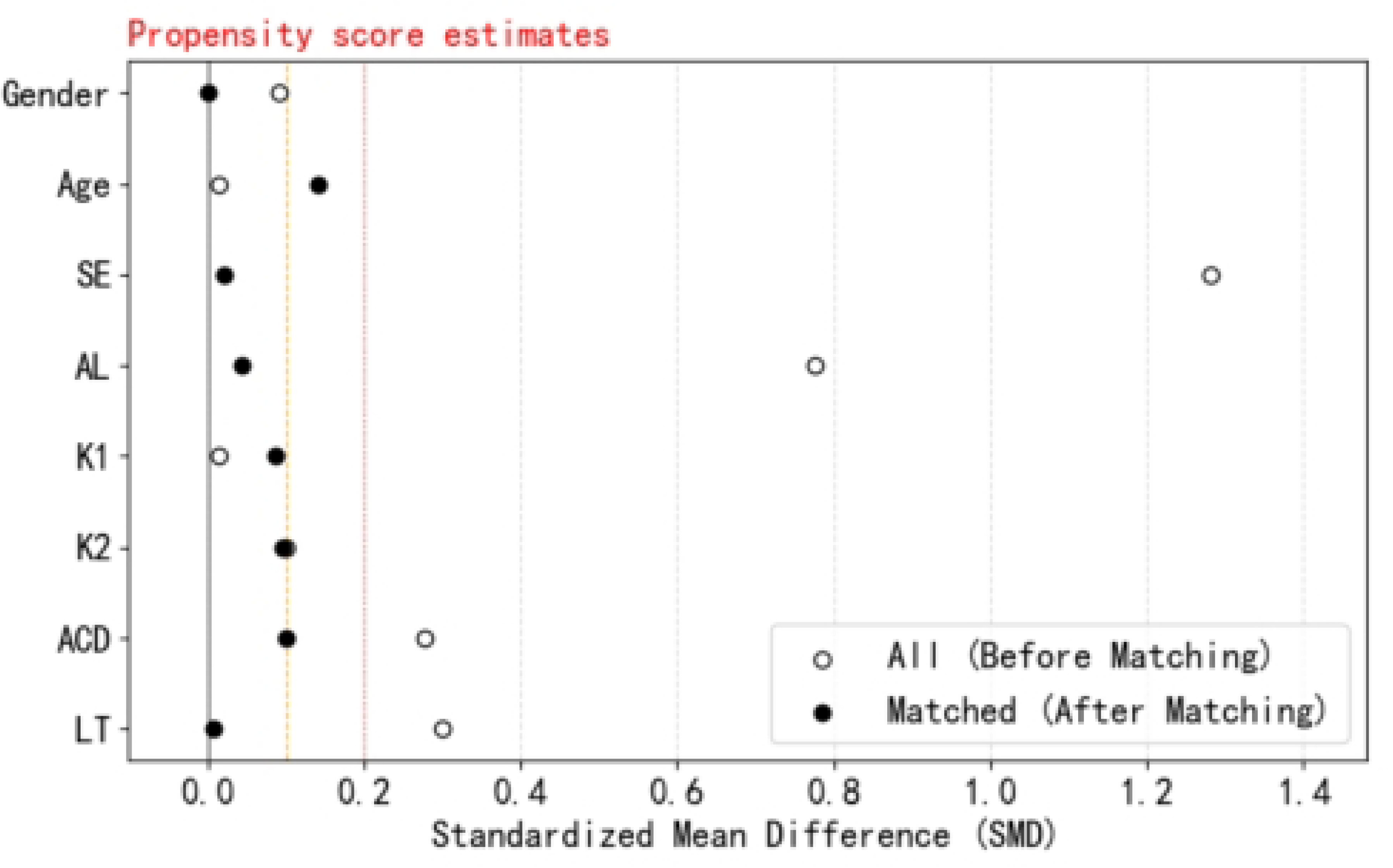
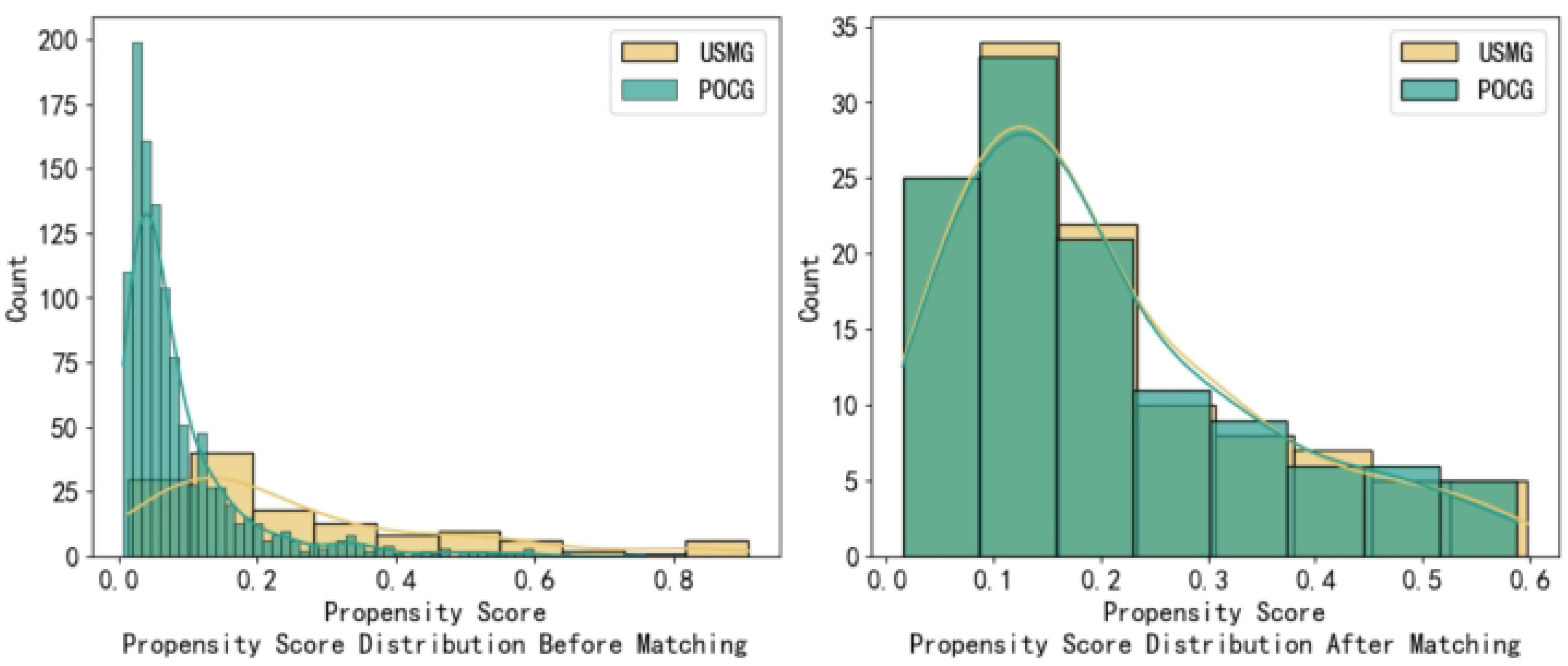
Love Plot and Distribution Fitting Curves for Baseline Characteristics Before and After Propensity Score Matching Between POCG and USMG.

**Table 2.** Balance assessment of baseline characteristics between the Under-Corrected Group(UCG) and Protocol-Defined Normal Vision Group(POCG) after propensity score matching(PSM)

| Variable<br>Name | Before Matching<br>( n=266/134 ) |  | \|SM<br>D\| | After Matching<br>( n=94/94 ) |  | \|SMD\| |
| --- | --- | --- | --- | --- | --- | --- |
|  | UCG | POCG |  | UCG | POCG |  |
| Male, n (%) | 71 (53.0%) | 545<br>(48.4%) | 2.64 | 59 (50.9%) | 59 (50.9%) | 0 |
| Age (years) | $9.50 \pm 0.27$ | $9.50 \pm 0.29$ | 0.01 | $9.50 \pm 0.27$ | $9.540 \pm 0.29$ | 0.04 |

| Variable<br>Name | Before Matching |  |  | After Matching |  |  |
| --- | --- | --- | --- | --- | --- | --- |
|  | ( n=266/134 ) |  | \|SM<br>D\| | ( n=94/94 ) |  | \|SMD<br>\| |
|  | UCG | POCG |  | UCG | POCG |  |
| CDVA(logMAR) | 0.19 ± 0.14 | 0.00 ± 0.02 | 1.64 | 0.18 ± 0.15 | 0.00 ± 0.02 | 1.72 |
| SE(D) | -2.43 ± 1.10 | -1.40 ± 0.77 | 1.64 | -2.16 ± 0.87 | -2.18 ± 0.97 | 0.19 |
| AL(mm) | 24.42 ± 0.59 | 23.93 ± 0.63 | 0.39 | 24.34 ± 0.56 | 24.37 ± 0.58 | 0.08 |
| K1(D) | 42.89 ± 1.42 | 42.87 ± 1.44 | 0.11 | 42.86 ± 1.45 | 42.74 ± 1.42 | 0.03 |
| K2(D) | 44.19 ± 1.54 | 44.03 ± 1.55 | 0.24 | 44.11 ± 1.58 | 43.96 ± 1.59 | 0.03 |
| ACD(mm) | 3.24 ± 0.24 | 3.18 ± 0.24 | 0.25 | 3.23 ± 0.25 | 3.25 ± 0.19 | 0.1 |
| LT(mm) | 3.41 ± 0.19 | 3.47 ± 0.21 | 0.04 | 3.41 ± 0.20 | 3.41 ± 0.17 | 0.01 |

CDVA, corrected distance visual acuity;SE,spherical equivalent;AL,axial length; SE,spherical equivalent; K,keratometry; ACD,anterior chamber depth;LD,lens thickness.

**Table 3.** Comparison of baseline characteristics balance between Under-corrected group (UCG) and Untreated Screened Myopia Group (USMG) after propensity score matching(PSM)

| Variable<br>Name | Before Matching |  |  | After Matching |  |  |
| --- | --- | --- | --- | --- | --- | --- |
|  | ( n=266/1125 ) |  | \|SMD\| | ( n=201/201 ) |  | \|SMD\| |
|  | UCG | USMG |  | UCG | USMG |  |
| Male, n (%) | 104 (39.1%) | 545 (48.4%) | 0.19 | 81 (40.3%) | 81 (40.3%) | 0 |
| Age (years) | 9.50 ± 0.27 | 9.50 ± 0.29 | 0.02 | 9.51 ± 0.28 | 9.49 ± 0.28 | 0.09 |
| SE(D) | -2.82 ± 0.97 | -1.40 ± 0.77 | 1.77 | -2.53 ± 0.88 | -2.48 ± 0.88 | 0.06 |
| AL(mm) | 24.35 ± 0.61 | 23.93 ± 0.63 | 0.67 | 24.27 ± 0.63 | 24.25 ± 0.60 | 0.04 |
| K1(D) | 43.23 ± 1.41 | 42.87 ± 1.44 | 0.25 | 43.15 ± 1.49 | 43.05 ± 1.32 | 0.07 |
| K2(D) | 44.57 ± 1.54 | 44.03 ± 1.55 | 0.35 | 44.46 ± 1.65 | 44.27 ± 1.39 | 0.13 |
| ACD(mm) | 3.23 ± 0.23 | 3.18 ± 0.24 | 0.24 | 3.23 ± 0.23 | 3.24 ± 0.22 | 0.06 |
| LT(mm) | 3.43 ± 0.18 | 3.47 ± 0.21 | 0.211 | 3.43 ± 0.19 | 3.43 ± 0.20 | 0.01 |

SE,spherical equivalent;AL,axial length; SE,spherical equivalent; K,keratometry; ACD,anterior chamber depth;LD,lens thickness.

**Table 4.** Comparison of baseline characteristics balance between Protocol-Defined Optimal Correction Group (POCG) and Untreated Screened Myopia Group (USMG) after propensity score matching(PSM)

| Variable<br>Name | Before Matching |  |  | After Matching |  |  |
| --- | --- | --- | --- | --- | --- | --- |
|  | ( n=134/1125 ) |  | \|SMD\| | ( n=94/94 ) |  | \|SMD\| |
|  | POCG | USMG |  | POCG | USMG |  |
| Male, n (%) | 71 (53.0%) | 545 (48.4%) | 0.09 | 59 (50.9%) | 59 (50.9%) | 0 |
| Age (years) | 9.50 ± 0.27 | 9.50 ± 0.29 | 0.02 | 9.50 ± 0.27 | 9.54 ± 0.29 | 0.14 |
| SE(D) | -2.43 ± 1.10 | -1.40 ± 0.77 | 1.28 | -2.16 ± 0.87 | -2.18 ± 0.97 | 0.02 |
| AL(mm) | 24.42 ± 0.59 | 23.93 ± 0.63 | 0.78 | 24.34 ± 0.56 | 24.37 ± 0.58 | 0.04 |
| K1(D) | 42.89 ± 1.42 | 42.87 ± 1.44 | 0.01 | 42.86 ± 1.45 | 42.74 ± 1.42 | 0.09 |
| K2(D) | 44.19 ± 1.54 | 44.03 ± 1.56 | 0.1 | 44.11 ± 1.58 | 43.96 ± 1.59 | 0.01 |
| ACD(mm) | 3.24 ± 0.24 | 3.18 ± 0.24 | 0.28 | 3.23 ± 0.25 | 3.25 ± 0.19 | 0.1 |
| LT(mm) | 3.41 ± 0.19 | 3.47 ± 0.21 | 0.3 | 3.41 ± 0.20 | 3.41 ± 0.17 | 0.01 |

SE,spherical equivalent;AL,axial length; SE,spherical equivalent; K,keratometry; ACD,anterior chamber depth;LD,lens thickness.

For the UCG versus POCG comparison, 94 matched pairs were formed (Table 2). Post-matching, all covariates achieved SMD < 0.1 except non-cycloplegic spherical equivalent (SMD = 0.19). Corrected distance visual acuity differed significantly between groups (UCG: 0.18 ± 0.15 logMAR; POCG: 0.00 ± 0.02 logMAR).

For UCG versus USMG, 201 matched pairs were established (Table 3). For POCG versus USMG, 94 matched pairs were established (Table 4). Both matches achieved SMD < 0.1 for all baseline covariates. Figures 3–4 show that most SMDs converged toward zero after matching, confirming effective balance.

### Primary Outcome: Comparison of Axial Length Growth

Following propensity score matching, balanced cohorts were established to compare annualized axial length changes (ΔAL, mm/year) across different correction status groups. The primary outcome was ΔAL over the one-year period from baseline (T0) to final follow-up (T2). To assess the temporal stability of this outcome, interim axial elongation rates during the first half-year (T0–T1; ΔAL1) and second half-year (T1–T2; ΔAL2) were also evaluated. Results are presented in Figure 5.

**Figure 5.**
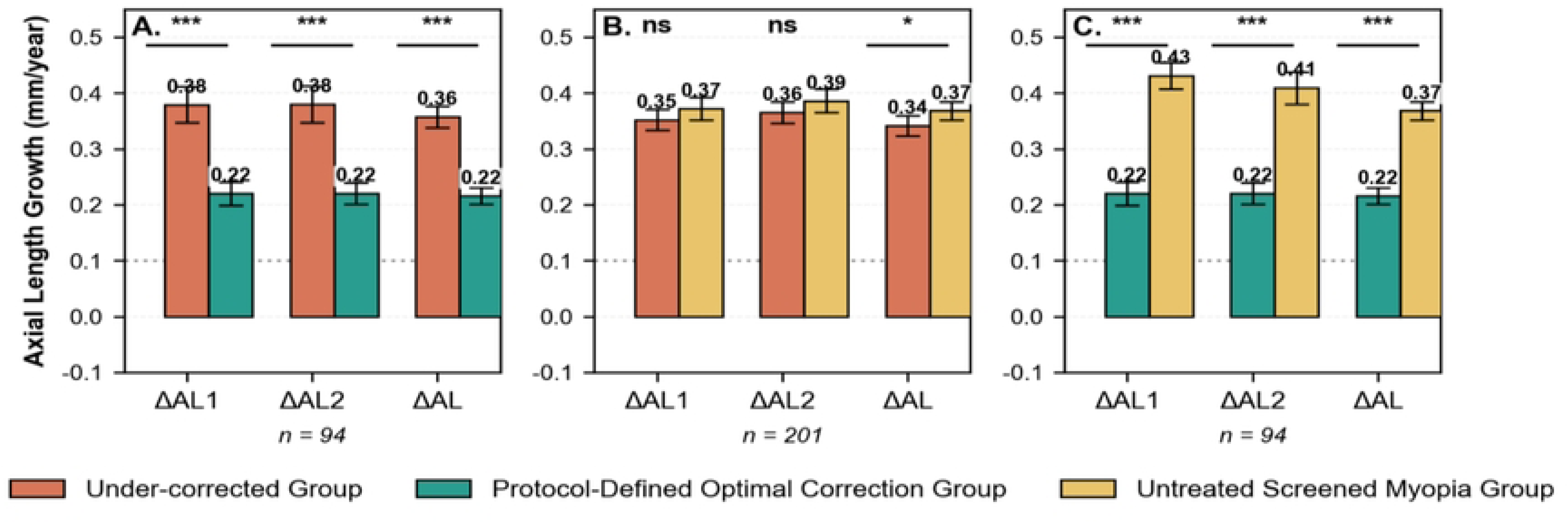
Annualized axial length growth in three comparison groups. *P<0.05, **P<0.01, ***P<0.001, ns: not significant.

Within spectacle wearers, POCG showed significantly lower ΔAL than UCG:ΔAL1 was 0.22 ± 0.20 vs. 0.37 ± 0.19 mm/year (P < 0.0001);ΔAL2 was 0.22 ± 0.18 vs. 0.38 ± 0.21 mm/year (P < 0.0001);Overall ΔALwas 0.23 ± 0.14 vs. 0.35 ± 0.14 mm/year (P < 0.0001).Across all time points, POCG demonstrated a 34.3% relative reduction in axial elongation (0.13 mm/year) compared to UCG(Table 5).

**Table 5.**
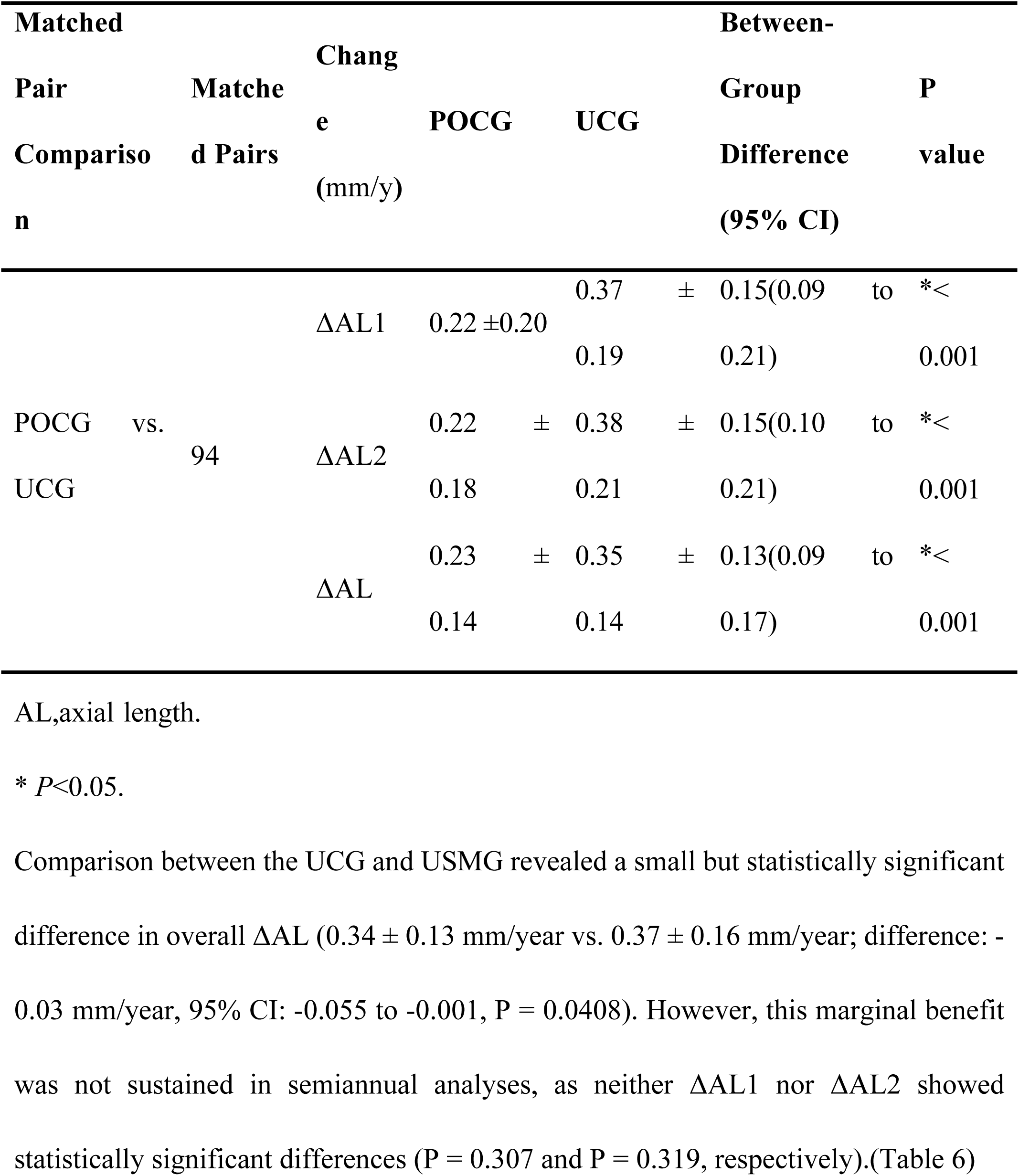
Comparison of Axial Length Change Outcomes between the Partial Optical Correction Group (POCG) and the Under-Correction Group (UCG) in Spectacle Wearers.

**Table 6.** Comparison of Axial Length Change between the Under-Correction Group (UCG) and the Untreated Screening Myopia Group (USMG)

| Matched |  | Change<br>(mm/y) | POCG |  | UCG |  | Between-Group<br>Difference<br>(95% CI) | P<br>value |
| --- | --- | --- | --- | --- | --- | --- | --- | --- |
| Pair<br>Comparison | Matched Pairs |  |  |  |  |  |  |  |
| UCG vs.<br>USMG | 201 | ΔAL1 | 0.35 | ± | 0.37 | ± | -0.02(-0.06 to 0.02) | 0.31 |
|  |  |  | 0.19 |  | 0.21 |  |  |  |
| | | ΔAL2 | 0.37 | ± | 0.39 | ± | -0.02(-0.06 to 0.02) | 0.32 |
|  |  |  | 0.21 |  | 0.21 |  |  |  |
| | | ΔAL | 0.34 | ± | 0.37 | ± | -0.03(-0.06 to 0.00) | *0.04 |
|  |  |  | 0.13 |  | 0.16 |  |  |  |

AL,axial length.
\* $P < 0.05$ .

**Table 7.**
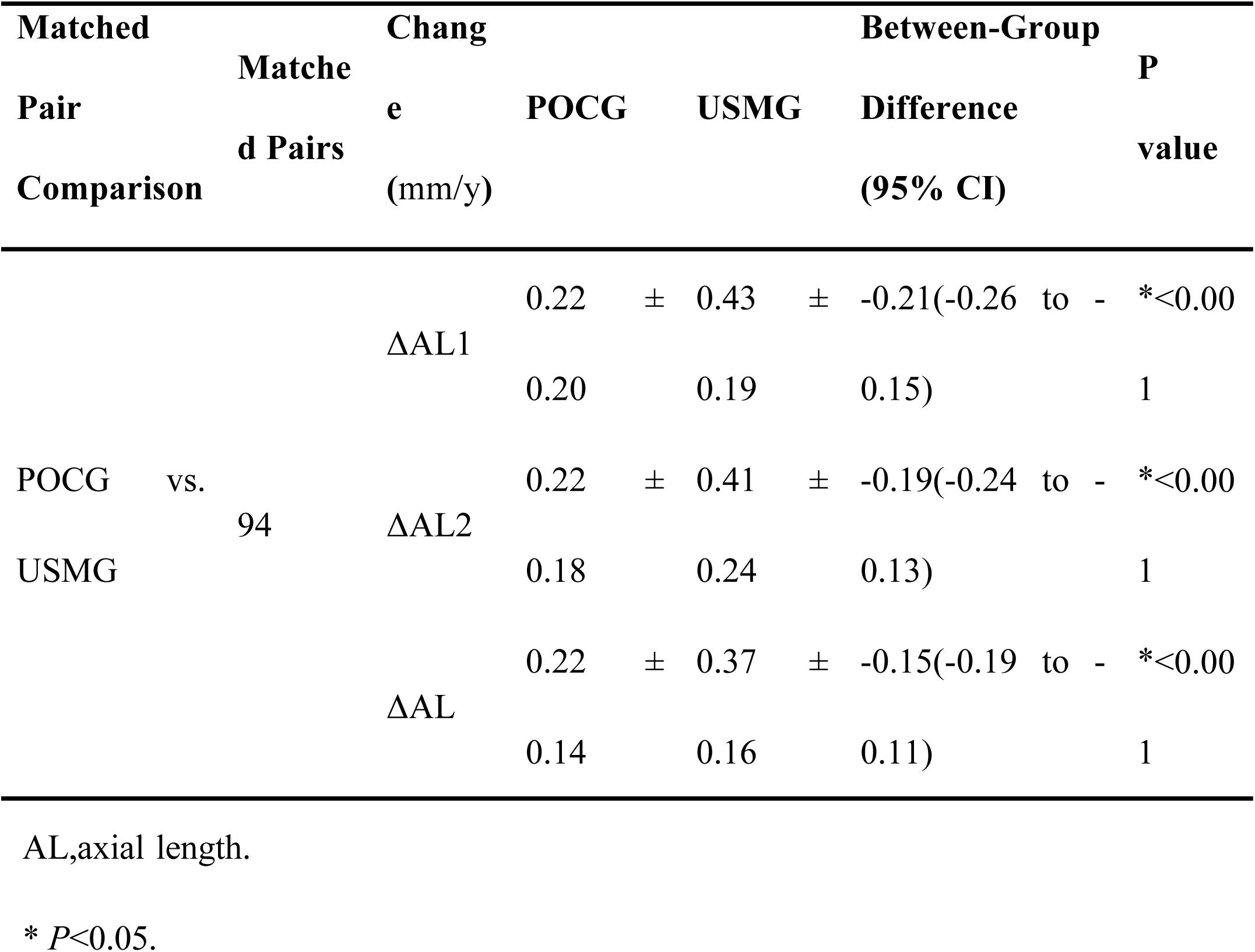
Comparison of Axial Length Change between the Partial Optical Correction Group (POCG) and the Untreated Screening Myopia Group (USMG)

## Discussion

This study, leveraging a real-world cohort from ordinary schools across China, systematically evaluated the impact of optical correction quality on myopia progression in children. Among spectacle-wearing children, sustained protocol-defined optimal correction (POCG) significantly attenuated annualized axial elongation compared to both the undercorrection group (UCG) and untreated screened myopia group (USMG), with reductions of 0.13 mm/year and 0.15 mm/year, respectively—differences of clear clinical significance. Secondary analyses further demonstrated that undercorrection (UCG) conferred only marginal and inconsistent protection relative to no correction (USMG): although statistically significant in annual pooled analyses, the clinical benefit was negligible and failed to persist in semiannual assessments.

We acknowledge that this observational cohort design cannot establish causality. Although a robust association exists between suboptimal corrected distance visual acuity and accelerated axial elongation, this relationship may reflect reverse causality (rapid progression inducing visual decline) or unmeasured confounding (e.g., spectacle wear compliance, near-work intensity, outdoor time). Critically, we do not assert that poor visual acuity causes accelerated progression; rather, we posit that regardless of its etiological role, compromised visual function may act as a critical amplifier within the progression cascade.

Specifically, even when visual impairment stems from rapid progression, chronic retinal defocus due to undercorrection may exacerbate axial elongation via a “blur–compensation–accelerated growth” vicious cycle. Insufficient correction often triggers excessive accommodation to achieve clarity, shifting the focal point posterior to the retina and intensifying hyperopic defocus—a well-established stimulus for ocular growth[16,17]. Optimal correction provides sharp foveal imaging, eliminating persistent hyperopic defocus while normalizing accommodative responses[16] and enhancing visual comfort[18], thereby attenuating aberrant optical signals driving axial elongation[19]. Consequently, functional visual status (CDVA) may serve dual roles: as a biomarker of progression and a modifiable intervention target.

Participants were recruited from ordinary schools in Chengdu rather than ophthalmic specialty clinics; hence, cycloplegic refraction was not performed. However, all subjects underwent identical non-cycloplegic examination protocols, ensuring that any measurement bias affected groups equally and preserving internal validity for intergroup comparisons. Future studies incorporating cycloplegic refraction would further validate these findings.

Notably, 73.77% of myopic children remained uncorrected (USMG) and 14.81% were undercorrected (UCG)—not a methodological flaw but a faithful reflection of real-world conditions across China and Asia. Prevailing cultural perceptions (e.g., viewing mild-to-moderate myopia as “requiring no intervention,” fearing spectacle dependency, or considering prescription as the endpoint of management) drive this pattern. While these perceptions limit generalizability to highly compliant cohorts, they underscore the study’s public health significance: we captured a large, overlooked population experiencing chronic subclinical visual blur in daily life—a potential environmental trigger for accelerated myopia progression.

Within the evidence hierarchy, ethical constraints preclude randomized assignment of children to prolonged undercorrection to assess harm. Thus, under current ethical frameworks, high-quality observational studies employing propensity score matching to control confounding represent the highest attainable evidence level for this clinical question. Our findings not only elucidate associations between real-world behaviors and health outcomes but also yield actionable insights directly translatable to policy and clinical practice.

Accordingly, we propose an actionable myopia control framework: (1) Transform vision screening from static snapshots to dynamic monitoring—shifting from 1–2 annual school screenings to quarterly or monthly high-frequency assessments by trained personnel using standardized digital platforms; (2) Implement a “prescription–verification–follow-up” closed-loop system—mandating visual function reassessment within 1–3 months post-prescription and quarterly CDVA target monitoring; (3) Redefine public health metrics—transitioning from process indicators (e.g., spectacle coverage) to outcome-oriented metrics (CDVA achievement rate, annualized axial elongation rate).

In conclusion, despite limitations inherent to observational design and non-cycloplegic refraction, rigorous statistical control and faithful characterization of real-world contexts reveal the foundational role of optical correction quality—particularly functional visual acuity—in myopia control. We advocate adopting clear vision as the ultimate criterion for intervention success, thereby bridging the critical gap between scientific evidence and tangible visual health benefits for children worldwide.

## Data Availability

Due to the sensitive nature of the data involving minors (9-year-old children) and privacy protection regulations approved by the Ethics Committee of the Affiliated Eye Hospital of Chengdu University of Traditional Chinese Medicine, the raw data cannot be made publicly available in a repository. However, de-identified data underlying the results presented in this study are available upon reasonable request. Qualified researchers may submit a data access request to the corresponding author, Dr. Junguo Duan, via email at . Requests will be reviewed by the Ethics Committee of the Affiliated Eye Hospital of Chengdu University of Traditional Chinese Medicine to ensure compliance with ethical standards and data privacy protocols before access is granted.

## Acknowledgements

The authors thank Bo Liu for his assistance with this study and all the participants for their participation.

## Notes

### Competing Interest Statement

The authors have declared no competing interest.

### Funding Statement

Yes

### Author Declarations

The study adhered to the tenets of the Declaration of Helsinki. Ethical approval was granted by the Ethics Committee of the Affiliated Eye Hospital of Chengdu University of Traditional Chinese Medicine (Approval No.: 2019YH-007). Written informed consent was obtained from parents or legal guardians of all participants through collaboration with the Chengdu Municipal Education Bureau.

